# Artificial intelligence-enabled echocardiography as a surrogate for multi-modality aortic stenosis imaging: post-hoc analysis of a clinical trial

**DOI:** 10.1101/2025.03.26.25324690

**Authors:** Evangelos K. Oikonomou, Neil J. Craig, Gregory Holste, Sumukh Vasisht Shankar, Audrey White, Menaka Mahendran, David E. Newby, Marc R. Dweck, Rohan Khera

## Abstract

**Background:** Accurate aortic stenosis (AS) phenotyping requires access to multimodality imaging which has limited availability. The Digital Aortic Stenosis Severity Index (DASSi), an AI biomarker of AS-related remodeling on 2D echocardiography, predicts AS progression independent of Doppler measurements. Whether DASSi-enhanced echocardiography provides a scalable alternative to multimodality AS imaging remains unknown. We sought to evaluate the ability of DASSi to define personalized AS progression profiles and validate its performance against multimodality imaging features of functional, structural, and biological disease severity.

**Methods:** In the SALTIRE-2 trial of participants with mild-or-moderate AS, we performed blinded DASSi measurements (probability of severe AS, 0-to-1) on baseline transthoracic echocardiograms. We evaluated the association between baseline DASSi and (i) disease severity by hemodynamic (peak aortic valve velocity [AV-V_max_]), structural (CT-derived aortic valve calcium score [AVCS]) and biological features ([18F]sodium fluoride [NaF] uptake on Positron Emission Tomography-CT), (ii) disease progression (change in AV-V_max_ and AVCS), and (iii) incident aortic valve replacement (AVR). We used generalized linear mixed, or Cox models adjusted for risk factors and aortic valve area, as appropriate.

**Results:** We analyzed 134 participants (72 [IQR: 69-78] years, 27 [20.1%] women) with a mean baseline DASSi of 0.51 (standard deviation [SD]: 0.19). DASSi was independently associated with disease severity: each SD increase was associated with higher AV-V_max_ (+0.21 [95%CI: 0.12-0.30] m/sec), AVCS (+284 [95%CI: 101-467] AU) and [18F]NaF TBR_max_ (+0.17 [95%CI: 0.04-0.31]). Higher DASSi was also associated with disease progression by Doppler (AV-V_max_) and CT (AVCS) at 24 months (*p*_interaction_ for DASSi (*x*) time<0.001), and future AVR (75 events over 5.5 [IQR: 2.4-7.2] years, adj.HR 1.47 [95%CI: 1.12-1.94] per SD).

**Conclusions:** DASSi is associated with functional, structural and biological features of AS severity as well as disease progression and outcomes. DASSi-enhanced echocardiography provides a readily accessible alternative to multimodality imaging of AS which has potential value both in clinical practice and as a clinical trial biomarker.

## INTRODUCTION

Aortic stenosis (AS) remains a prevalent condition without an effective medical therapy.^1,2^ Several medical interventions targeting a broad range of pathophysiological processes have repeatedly failed to prevent AS progression in randomized clinical trials (RCTs).^3–6^ Nevertheless, recent discoveries on the causal role of lipoprotein (a) (Lp(a)) in aortic valve disease,^7–9^ and the development of targeted therapeutics,^10–12^ have generated a growing interest in optimizing the design of trials in AS to balance efficiency and scalability with precision phenotyping. Trials in early AS remain limited by long development and testing cycles due to relatively slow progression rates.^13^ Doppler-defined features do not accurately identify progression risk,^13,14^ suggesting a need for multimodality imaging of functional, structural, and biological features of AS severity.^2,15–20^ This includes computed tomography (CT) imaging to quantify the aortic valve calcium burden,^6,21^ cardiac magnetic resonance imaging (CMR) to quantify myocardial injury,^22^ and molecular imaging with targeted tracers such as [18F]sodium fluoride ([18F]NaF) to examine microcalcification activity.^23^ Unfortunately, these modalities are not readily available across many sites, limiting their use across large, multi-center clinical studies evaluating novel medical therapies. Together, these challenges increase the financial and time investment required to evaluate potential new medical therapies.

Digital aortic stenosis severity index (DASSi) is a recently described and validated AI-enabled biomarker of AS derived from single-view 2D echocardiography of the parasternal long axis (PLAX) view.^14,24^ DASSi was trained to identify valvular and myocardial remodeling patterns of severe AS from PLAX videos, thus optimized for scalable use across standard and portable acquisitions. It provides a continuous score ranging from 0 to 1 that can discriminate the presence of AS and its severity stages,^24^ whilst improving the prediction of future AS progression across multi-center retrospective analyses and independent of Doppler measures.^14^ Given its simplicity, DASSi offers a scalable approach to the phenotyping of aortic sclerosis and early stages of AS using rapid and observer-independent protocols. However, external validation in a protocolized setting free of referral bias, and multimodality validation against functional, structural, and biological correlates derived from multimodality imaging represent essential knowledge to understand whether DASSi-enhanced echocardiography may provide an alternative strategy to multimodality imaging in clinical trials of AS therapies and the phenotyping of patients with non-severe AS.

To address these key questions, we present a comprehensive evaluation of DASSi against multiparametric phenotyping of the natural history of mild and moderate AS progression in a cohort of individuals undergoing protocolized follow-up by serial multimodality imaging. In a *post hoc* analysis of the SALTIRE-2 (Study Investigating the Effect of Drugs Used to Treat Osteoporosis on the Progression of Calcific Aortic Stenosis 2) randomized clinical trial, one of the key clinical trials deploying multimodality imaging to explore functional, structural and molecular changes in AS-related remodeling over 24 months,^6^ we applied DASSi in a blinded fashion and examined its association with cross-sectional features of functional, structural, and biological AS severity, longitudinal disease progression by hemodynamic severity and aortic valve calcification, and clinical outcomes. This approach aims to evaluate a paradigm of using AI-enhanced echocardiographic biomarkers as a scalable and accessible alternative to multimodality imaging to support the design of future clinical trials for medical therapies in AS.

## METHODS

### Data Source and Patient Population

#### Data source

We designed a study to examine the cross-sectional and prognostic implications of DASSi phenotyping among patients with mild or moderate AS undergoing protocolized multimodality imaging using data from the SALTIRE-2 trial (NCT02132026, Scotland A Research Ethics Committee, 14/SS/0064).^6^ Briefly, SALTIRE-2 was a single-center, parallel-group, double-blind, randomized clinical trial that enrolled patients >50 years of age with mild or moderate aortic stenosis at baseline, as defined by a peak aortic valve velocity (AV-V_max_) of >2.5 m/sec on TTE and grade II-IV aortic valve calcification. Participants were identified from cardiology outpatient clinics across South-East Scotland. Notable exclusion criteria included anticipated or planned aortic valve surgery in the next 6 months, life expectancy < 2 years, long-term corticosteroid use, inability to stand or sit for more than 30 minutes, major or untreated cancers, women of childbearing potential, chronic kidney disease with an estimated glomerular filtration rate of <30 mL/min/1.73 m^2^, and allergy or contraindication to iodinated contrast. A complete list of inclusion and exclusion criteria has been previously published.^6^

#### Study exposure and outcomes

In line with the original trial protocol,^6^ participants underwent baseline TTE and combined [18F]NaF PET-CT and non-contrast CT followed by randomization to denosumab, placebo injection, oral alendronic acid, or matching placebo capsule (2:1:2:1 ratio). Participants were followed by echocardiography every 6 months (6, 12, 18, 24 months), with repeat combined [18F]NaF PET-CT and non-contrast CT performed at 12 months and non-contrast CT alone at 24 months.

### Study Aims

Our analysis was designed with three key aims. In Aim 1, we examined the association between baseline DASSi (computed from pre-randomization TTE studies) and disease severity: baseline multimodality imaging-derived features of TTE-derived Doppler parameters, aortic valve calcium score (AVCS) by CT and active microcalcification by [18F]NaF uptake on PET-CT. In Aim 2, we further evaluated the links between DASSi and future disease progression by hemodynamic (AV-V_max_) and structural (AVCS) features over 24 months (Aim 2). Finally, we examined the association between baseline DASSi and the incidence of adverse clinical outcomes, namely AVR and all-cause mortality (Aim 3).

### DASSi Phenotyping

DASSi represents a digital echocardiographic biomarker derived from a convolutional neural network applied to 2D-echocardiographic acquisitions of the PLAX view that has been trained to provide a quantitative metric of the probability of AS (ranging from 0 [lowest probability] to 1 [highest probability]).^14,24^ For reference, values of 0.607 or greater may be used to screen for severe AS, although a dose-response relationship exists between DASSi and cross-sectional severity of AS, ranging from early sclerosis to severe AS.^14,24^ DASSi was computed across available baseline echocardiograms in a blinded fashion through an executable software application that automated all sequential steps of deidentification, image down-sampling, automated view classification of PLAX videos, and inference. Study-level DASSi estimates represent the mean of video-level predicted probabilities. This ensured data privacy without the transfer of images across sites.

### Outcome assessment

#### Functional (hemodynamic) measures by Doppler echocardiography

Functional metrics of AS severity were derived from Doppler echocardiography in line with international guidelines. As per the original report,^6^ standard 2-dimensional views and pulsed and continuous wave Doppler measurements were acquired with measurements averaged over 3 cardiac cycles, or 5 cycles if in atrial fibrillation. Aortic valve mean gradient (in mm Hg) was derived from the Bernoulli equation, whereas aortic valve area (AVA, in cm^2^) was estimated using the continuity equation.^18^ These parameters represent standard metrics of the echocardiographic assessment of AS and are central to defining the severity stage by guidelines.^18^ The detailed imaging analysis protocol has been previously published.^6^

#### Structural (CT) and biological activity ([18F]NaF PET-CT) outcomes

To define the structural severity of AS we quantified the AVCS on non-contrast CT scans using a standardized technique,^25,26^ with regions of interest (ROI) drawn around areas of valvular calcification on sequential axial slices, and a standard threshold of 130 Hounsfield units. Following segmentation, the Agatston score (Agatston units, AU) was semi-automatically calculated using standard weightings.^27^ As a surrogate of biological disease activity, we measured [18F]NaF aortic valve uptake as described previously, ^28,29^ using a standardized method with a 6-mm height polyhedron centered, on the valvular region of highest valvular uptake in the z-plane and contoured manually around the valve perimeter to calculate mean and maximum standardized uptake values. The mean and maximum target-to-background ratios (TBR) were calculated by dividing the mean and maximum standardized uptake values in the region of interest by the mean blood pool standardized uptake value calculated from a 2-cm^2^ region drawn in the center of the right atrium at the level of the right coronary ostium.

#### Clinical outcomes

Participants were followed prospectively for the incidence of AVR (surgical or transcatheter) and all-cause mortality through study interviews, as well as a review of their electronic health records. Study enrollment was performed between August 19, 2015, and November 6, 2017 with the last visit in November, 2019. Clinical outcomes were collected up until August 2024.

### Statistical Analysis

We performed pooled analyses of all eligible individuals, recognizing the lack of treatment effect of either alendronic acid or denosumab on the study’s primary and secondary outcomes.^6^ Categorical values are summarized as counts (percentages), and continuous variables are summarized as mean (standard deviation [SD]) or median (25^th^-75^th^ percentile). Bivariate correlations between continuous variables are presented using the Spearman’s rho (ρ) correlation coefficient.

In Aim 1, the cross-sectional association between baseline DASSi and imaging parameters of functional severity (AV-V_max_, aortic valve mean gradient, AVA), structural (AVCS) severity, or biological activity ([18F]NaF mean and maximal target-to-background ratio [TBR_mean_ and TBR_max_]) was examined in generalized linear models with each one of these metrics as the dependent variable, and DASSi, age, sex, hypertension, diabetes mellitus, smoking status, total cholesterol and systolic blood pressure as the independent covariates. For structural (CT-derived AVCS) and biological (PET-CT-derived) features, Doppler-derived AVA was included as an additional independent covariate to explore the Doppler-independent association of DASSi with these multimodality imaging-derived metrics. The goodness-of-fit of nested models with and without the addition of DASSi was compared by the likelihood ratio (LR) test. For visualization purposes, we present both box-and-whisker plots of observed values across distinct DASSi groups and fitted lines from multivariable spline regression models with k=3 knots.

In Aim 2, we explored the independent association of DASSi with longitudinal changes in both functional parameters by serial Doppler echocardiography (AV-V_max_) and structural severity by CT imaging (AVCS). We fit generalized mixed linear models with each one of these metrics as the dependent variable, and DASSi at baseline, time (in months), the interaction between the two (baseline DASSi *x* time), age, sex, hypertension, diabetes mellitus, smoking status, total cholesterol, systolic blood pressure, denosumab or alendronic acid use, and the baseline AVA as independent covariates. Including the interaction term allowed us to examine the hypothesis that higher DASSi at baseline is associated with faster progression in AS severity. Furthermore, given the clustering of observations across visits of the same participant, we included participant as a random effect. We summarize the results of the models by plotting the adjusted AV-V_max_ and AVCS across discrete levels of baseline DASSi.

In Aim 3, we further evaluated the association between DASSi and clinical outcomes, such as AVR, with all-cause mortality also presented for reference. Here, we present adjusted survival curves (using the same covariates as outlined above) derived from multivariable Cox regression models for time-to-AVR. To provide insights into discrimination, we report the C-statistic and delta C-statistic for different models accompanied by 95% confidence intervals derived from bootstrapping with 1000 replications.

Finally, we evaluated the relative change in the required sample size for a hypothetical study with a DASSi-led enrollment strategy enriching for the highest-risk group (≥0.7) versus an AV-V_max_-guided enrollment (≥3 m/sec) approach. For each one of these two scenarios, we applied paired one-sample t-test power calculations (alpha 0.05, power 0.9) to detect a significant change in AV-V_max_ at 1 year, based on the observed (mean, standard deviation) of annualized AV-V_max_ change across these subgroups in the SALTIRE-2 population.^30^

Across all analysis, alpha (α) was set at 0.05 with no adjustment for multiple comparisons. All analyses were performed using Python version 3.11.2 (Python Software Foundation) and R version 4.2.3 (R Foundation) statistical software. As a post hoc observational analysis of a clinical trial evaluating DASSi as a clinical biomarker, reporting stands consistent with the STROBE (Strengthening the Reporting of Observational Studies in Epidemiology) statement.^31^

## RESULTS

### Patient Population

Out of 150 participants in the original SALTIRE-2 trial, a total of 134 (89.3%) individuals underwent successful DASSi phenotyping at baseline and were included in this analysis (n=2 had no available images, and in n=14 did not pass DASSi-specific quality control) (**Figure 1**). Among these 134 participants, a total of 44 (32.8%) individuals were randomized to alendronic acid, 46 (34.3%) to denosumab and the remaining 44 (32.8%) to matching placebo (injection or capsules). The median age was 72 [IQR: 68-78] years, and 27 (20.1%) were women (**Table 1**).

**Figure 1.**
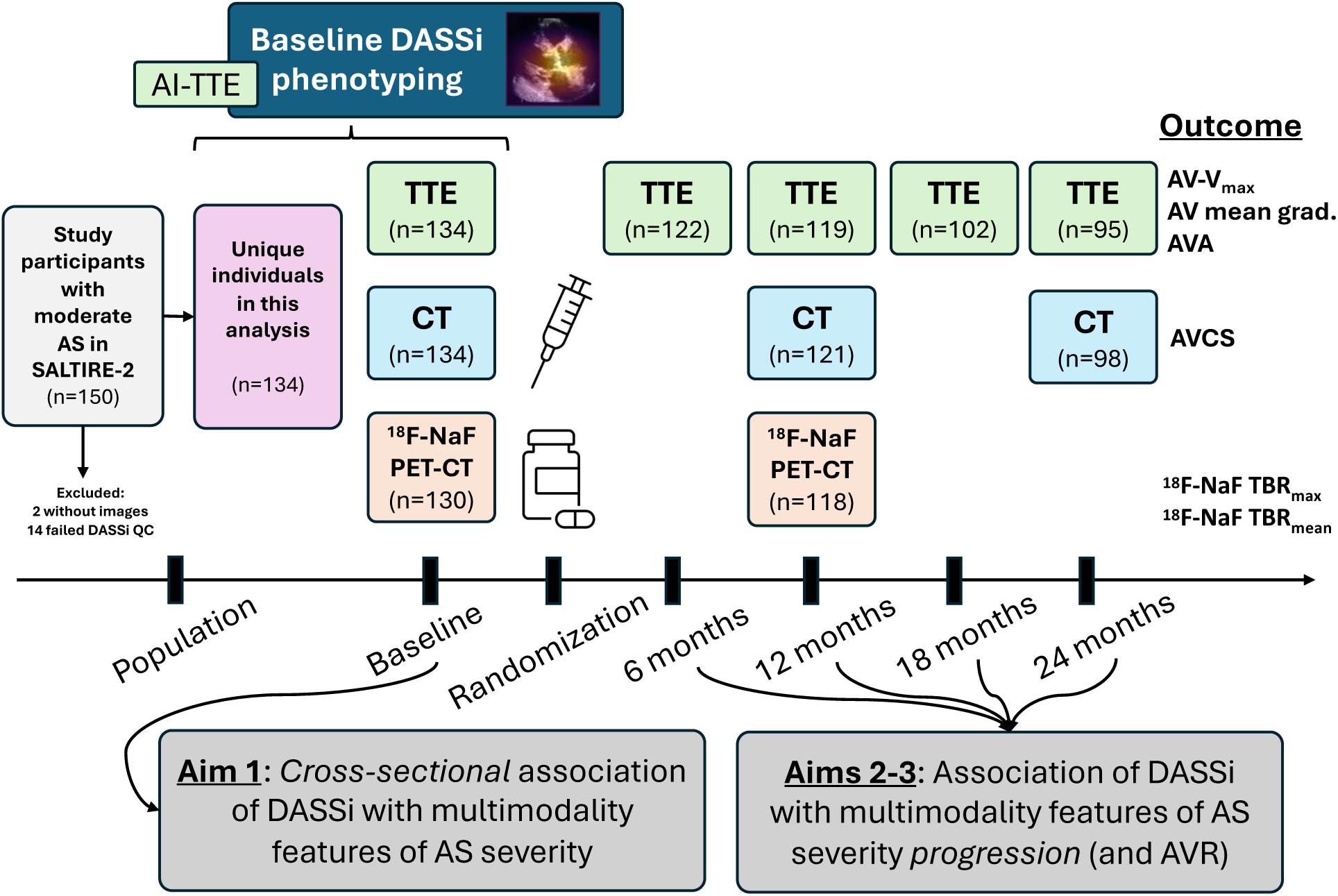
Overview of the study protocol. We performed a post hoc analysis of 134 individuals who were included in the original SALTIRE-2 trial and were randomized to treatment with denosumab vs alendronic acid vs placebo. Participants had baselined transthoracic echocardiography (TTE) demonstrating mild to moderate aortic stenosis (AS) and were followed up over 2 years (24 months) by serial TTE (0, 6, 12, 18, 24 months), CT (computed tomography) imaging to quantify the aortic valve calcium score (AVCS, 0, 12 and 24 months) as well as [18F]NaF-PET-CT (positron emission tomography CT) imaging (0 and 12 months) to assess aortic valve microcalcification. DASSi measurements were performed at baseline and were subsequently linked to cross-sectional and longitudinal patterns of AS severity and progression. *AI: artificial intelligence; AS: aortic stenosis; AV: aortic valve; AVCS: aortic valve calcium score; AV- V_max_: peak aortic valve velocity; AVA: aortic valve area; CT: computed tomography; DASSi: digital aortic stenosis severity index; NaF: sodium fluoride; PET: positron emission tomography; TBR: target-to-background ratio; QC: quality control*.

**Table 1.** Study population characteristics.

|  | Overall |
| --- | --- |
| **Total participants** | 134 (100) |
| **Age (years)** | 72 [68, 78] |
| **Sex** |  |
| Male | 107 (79.9) |
| Female | 27 (20.1) |
| **Ethnicity** |  |
| Scottish | 121 (90.3) |
| Irish | 1 (0.7) |
| Other British | 10 (7.5) |
| Other White | 2 (1.5) |
| **Risk Factors** |  |
| Hypertension | 101 (75.4) |
| Diabetes mellitus | 31 (23.) |
| Total cholesterol (mmoL/L) | 4.2 [3.6, 5.0] |
| **Smoking** |  |
| Current | 12 (9.0) |
| Former | 62 (46.3) |
| **Medications** |  |
| ACEi | 51 (38.1) |
| ARB | 27 (20.1) |
| Beta-blocker | 50 (37.3) |
| Calcium channel blocker | 42 (31.3) |
| Thiazide diuretic | 26 (18.7) |
| Mineralocorticoid antagonists | 7 (5.2) |
| Alpha-1 blocker | 11 (8.2) |
| **Randomization drug** |  |
| Alendronic acid | 44 (32.8) |
| Denosumab | 46 (34.3) |
| **Baseline imaging** |  |
| ***Transthoracic Doppler Echocardiography*** |  |
| AV-V<sub>max</sub> (m/sec) | 3.4 [3.0,3.8] |
| AV mean gradient (mmHg) | 24.0 [18.3,32.1] |
| AVA (cm<sup>2</sup>) | 1.1 [0.9,1.2] |
| Digital Aortic Stenosis Severity index (DASSi) | 0.51 (0.19) |
| ***Computed tomography (CT) imaging*** |  |
| AV calcium score (Agatston Units, AU) | 1158 [620, 2064] |
| ***Positron Emission Tomography (PET) imaging*** |  |
| [18F]NaF TBR (mean) | 1.6 [1.4, 1.8] |
| [18F]NaF TBR (max) | 2.6 [2.2, 3.0] |

Data presented as counts [percentages] and mean (standard deviation) or median [interquartile range], as appropriate. ACEi: angiotensin-converting enzyme inhibitor; ARB: angiotensin II-receptor blocker; AV: aortic valve; AV-V<sub>max</sub>: peak aortic valve velocity; AVA: aortic valve area; CT: computed tomography; DASSi: digital aortic stenosis severity index; NaF: sodium fluoride; PET: positron emission tomography; TBR: target-to-background ratio.

All 134 participants had Doppler echocardiography and CT imaging to quantify AVCS at baseline, and 130 (97.0%) had available [18F]NaF PET-CT data, with 95 (70.9%) and 98 (73.1%) patients undergoing a TTE and CT scan at 24 months respectively (**Figure 1**). The median AV-V_max_ at baseline was 3.4 [IQR: 3.0-3.8] m/sec, with a median aortic valve gradient of 24.0 [IQR: 18.3-32.1] mm Hg and a median calculated AVA of 1.1 [IQR: 0.9-1.2] cm^2^. On CT imaging the median AVCS was 1,158 AU (Agatston Units) [IQR: 620-2064] (n=134 [100%]).

### Baseline DASSi and functional, structural and biological features of AS

The mean DASSi at baseline was 0.51 (SD: 0.19). Cross-sectionally, higher baseline DASSi was independently associated with worse functional (hemodynamic), structural and biological metrics of AS severity and disease activity. Each SD higher DASSi was associated with 0.21 [95%CI: 0.12-0.30] m/sec higher adjusted AV-V_max_, 3.68 [95% CI: 2.18-5.19] mm Hg higher mean aortic valve gradient, and a 0.07 [0.03-0.12] cm^2^ lower AVA (**Supplemental Table 1**). On CT and PET-CT-imaging, each SD increment in DASSi was further associated with a 284 [95%CI: 101- 467] AU greater CT-derived AVCS, and a 0.08 [95%CI: 0.03-0.13] and 0.17 [95%CI: 0.04-0.31] increase in [18F]NaF TBR_mean_ and TBR_max_, independent of clinical risk factors and Doppler-defined AVA. Incorporating DASSi to a baseline model consisting of age, sex, and AVA increased the model’s goodness-of-fit and performance both for CT-derived AVCS and PET-CT- derived TBR_max_ (*p*<0.001 by likelihood ratio test for both; **Figure 2** and **Supplemental Figure 1**).

**Figure 2.**
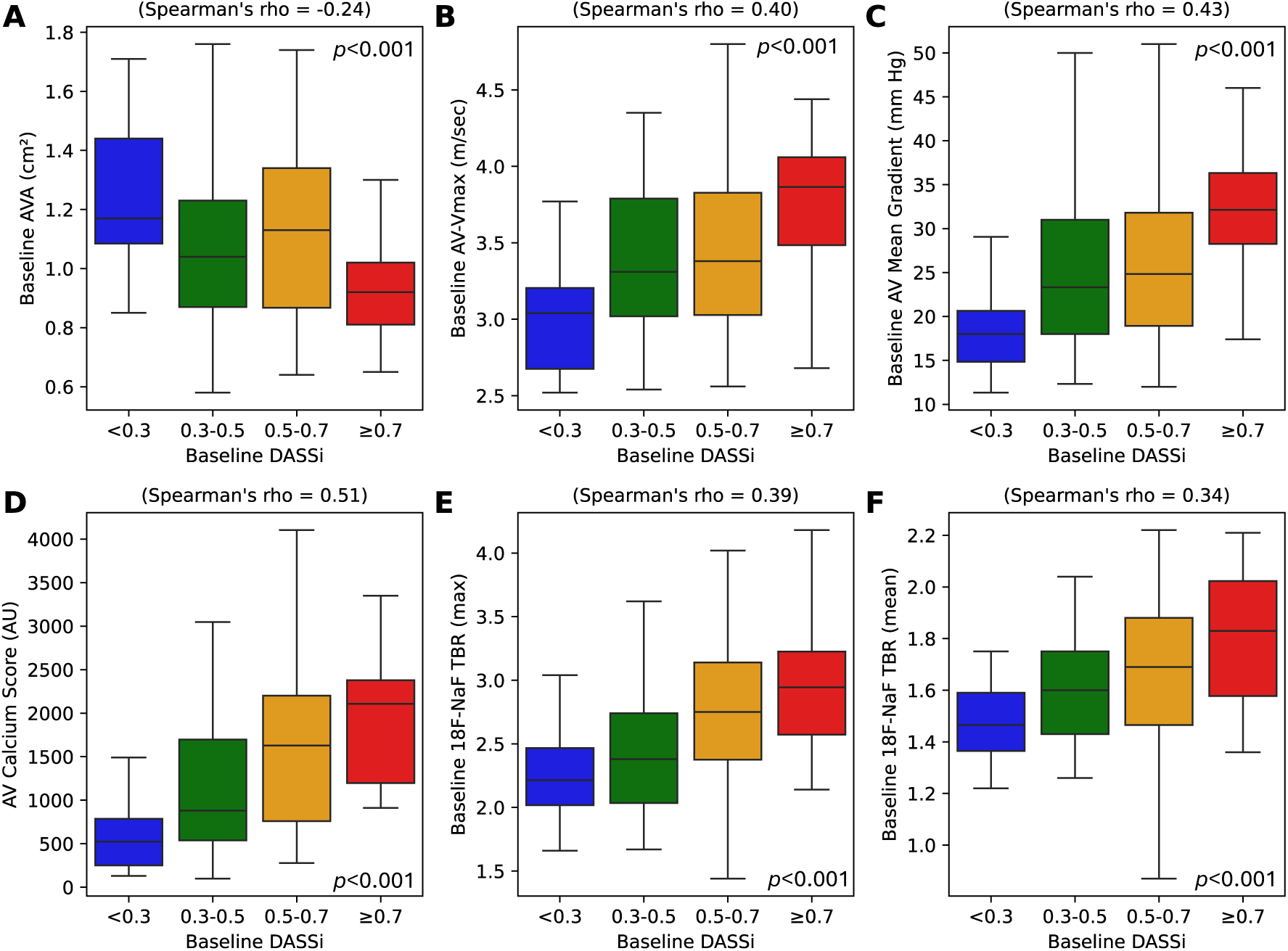
Cross-sectional correlation of DASSi with multimodality AS phenotyping. Box and whisker plots summarizing the observed distribution of Doppler echocardiography metrics of AS severity (**A**: AVA, **B**: AV-V_max_, **C**: AV mean gradient), CT-derived aortic valve calcium score (AVCS, **D**), and [18F]NaF-PET-CT-derived metrics of aortic valve calcification (**E**: TBR_max_; **F**: TBR_mean_). For visualization purposes DASSi is discretized into four severity groups (<0.3 [blue]; 0.3 to <0.5 [green], 0.5 to <0.7 [orange]; ≥0.7 [red]). *AS: aortic stenosis; AV: aortic valve; AV-V_max_: peak aortic valve velocity; AVA: aortic valve area; AVCS: aortic valve calcium score; AU: Agatston Units; CT: computed tomography; DASSi: digital aortic stenosis severity index; NaF: sodium fluoride; PET: positron emission tomography; TBR: target-to-background ratio*.

### Baseline DASSi and disease progression in functional and structural AS features

#### Functional (hemodynamic) progression

Higher baseline DASSi was associated with faster progression in Doppler-derived parameters of hemodynamic severity by AV-V_max_, as evidenced by a positive interaction between baseline DASSi and time (**Table 2**). When indexed to the baseline visit, AV-V_max_ increased by 0.65 [95%CI 0.62-0.69] m/sec in the DASSi ≥0.7 group versus 0.28 [95%CI: 0.23-0.34] m/sec in the <0.3 group, with evidence of a dose-response relationship (**Figure 3**).

**Figure 3.**
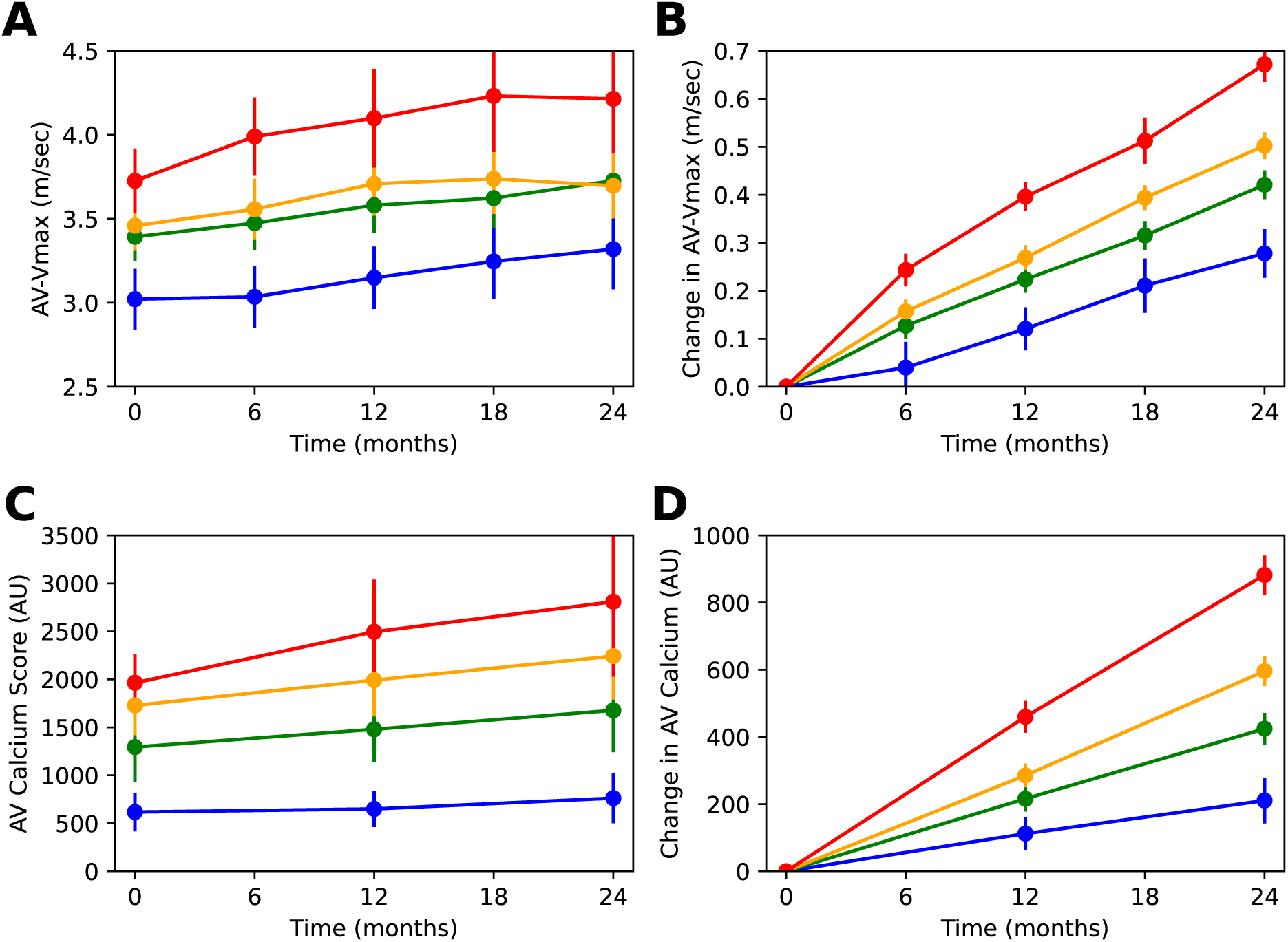
Baseline DASSi and AS progression by Doppler echocardiography and CT imaging. **(A)** Adjusted peak aortic valve velocity and (**C**) CT-derived aortic valve calcium score (AVCS) across time and study visits stratified by baseline DASSi groups (<0.3 [blue]; 0.3 to <0.5 [green], 0.5 to <0.7 [orange]; ≥0.7 [red]). (**B, D**) Adjusted absolute change in peak aortic valve velocity (AV-V_max_) and AVCS relative to baseline across time and study visits stratified by baseline DASSi groups. Point estimates are accompanied by 95% confidence intervals of the mean. *AS: aortic stenosis; AV: aortic valve; AV-V_max_: peak aortic valve velocity; AVCS: aortic valve calcium score; AU: Agatston Units; CT: computed tomography; DASSi: digital aortic stenosis severity index*.

**Table 2.** Association between baseline DASSi and longitudinal changes in multimodality AS severity metrics.

| Modality | Dependent Variable | Independent variable | Coefficient | 95% CI Lower | 95% CI Upper | *p* value |
| --- | --- | --- | --- | --- | --- | --- |
| TTE | AV-$V_{\max}$ (m/sec) | DASSi | 0.185 | 0.101 | 0.269 | <0.001 |
|  |  | DASSi x month | 0.002 | 0.000 | 0.013 | 0.034 |
|  | AV mean grad. (mm Hg) | DASSi | 3.35 | 1.86 | 4.84 | <0.001 |
|  |  | DASSi x month | 0.08 | 0.03 | 0.13 | 0.002 |
| CT | AVCS (AU) | DASSi | 305.61 | 116.42 | 494.79 | 0.002 |
|  |  | DASSi x month | 8.26 | 4.09 | 12.42 | <0.001 |

The estimates are derived from mixed linear models with DASSi at baseline, time (in months), the interaction between the two (baseline DASSi x time), age, sex, hypertension, diabetes mellitus, smoking status, total cholesterol, systolic blood pressure, denosumab or alendronic acid use, and baseline AVA (for AVCS) included as fixed effects, with participant included as a random effect. *AS*: aortic stenosis; *AU*: Agatston Units; *AV*: aortic valve; *AV- $V_{\max}$* : peak aortic valve velocity; *AVA*: aortic valve area; *AVCS*: aortic valve calcium score; *CI*: confidence interval; *CT*: computed tomography; *DASSi*: digital aortic stenosis severity index.

#### Structural progression

Higher baseline DASSi was also associated with greater progression in the structural severity by CT-derived AVCS over 24 months, independent of baseline Doppler parameters. Similarly to the observed progression in Doppler severity, the marginal increase in AVCS over 24 months was 867 [95%CI: 805-929] AU among those with DASSi ≥0.7 versus 220 [138-302] AU among those with DASSi <0.3 (**Figure 3**), with a significant interaction between baseline DASSi and time (**Table 2**).

### Baseline DASSi and future clinical events

Over a median follow-up of 5.5 [IQR: 2.4-7.6] years, 75 (56.0%) individuals underwent AVR, and 43 (32.1%) died. In multivariable analyses adjusting for age, sex, baseline cardiovascular risk factors, as well as baseline AVA, higher baseline DASSi levels were associated with a higher risk of AVR (per SD increment in DASSi: adj. HR 1.47 [95%CI: 1.12-1.94], *p*<0.001), with an attenuated and non-significant effect seen for all-cause mortality (adj. HR 1.21 [95%CI: 0.83-1.77]). Those in the highest DASSi stratum at baseline (≥0.7) had a nearly 3.3-fold higher adjusted risk of AVR (adj. HR 3.29 [95%CI: 1.06-9.61], *p*=0.039) when compared with the reference DASSi<0.3 group after adjusting for baseline AVA and risk factors (**Figure 4**).

**Figure 4.**
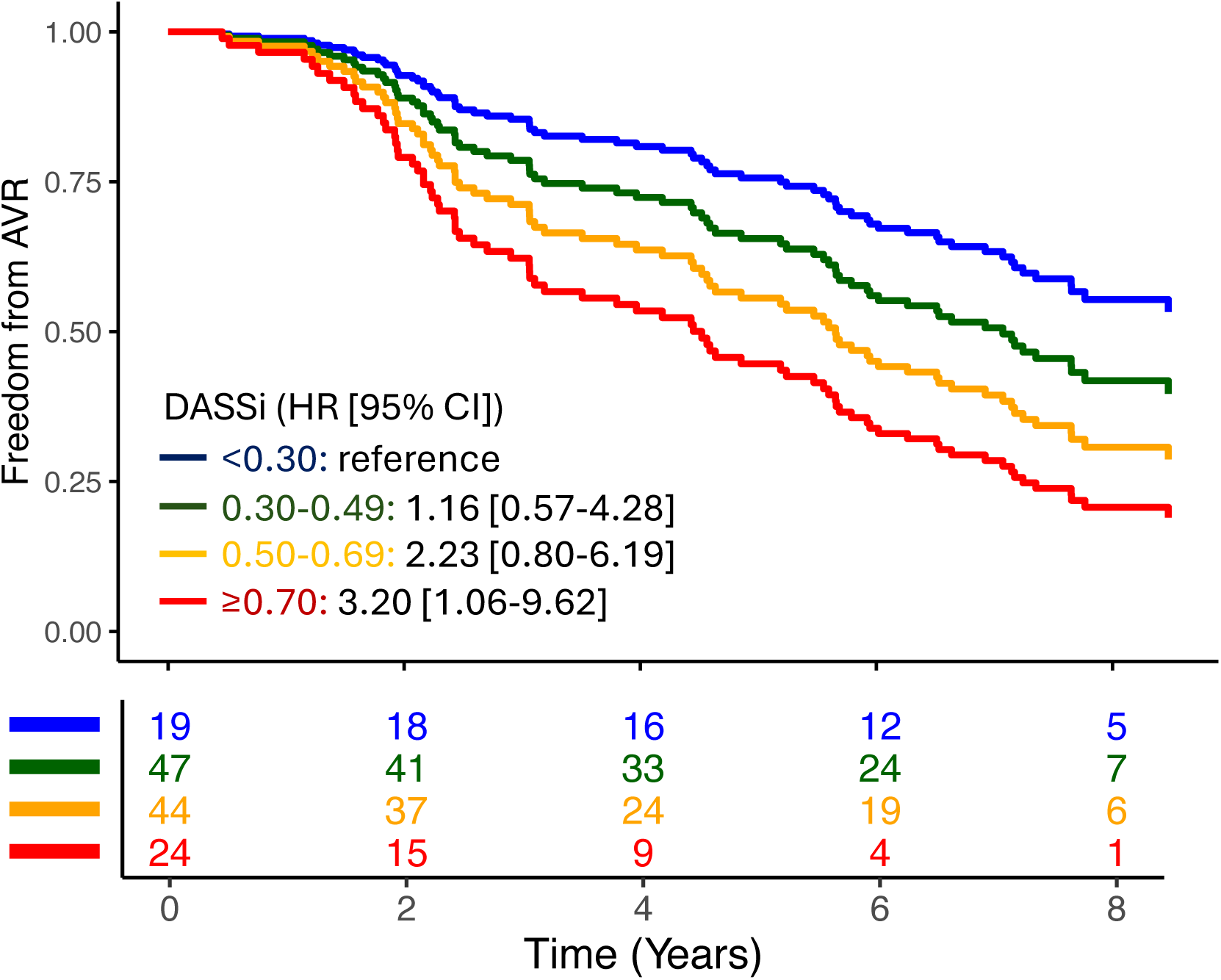
Baseline DASSi and incident aortic valve replacement (AVR). Adjusted survival curves for time-to-AVR stratified by baseline DASSi groups (<0.3 [blue]; 0.3 to <0.5 [green], 0.5 to <0.7 [orange]; ≥0.7 [red]). The figure is accompanied by a table of numbers at risk across 2-year intervals. *AVR: aortic valve replacement; CI: confidence interval; DASSi: digital aortic stenosis severity index; HR: hazard ratio*.

Of note, the C-statistic for a baseline model incorporating Doppler-derived AVA along with demographics and clinical risk factors (age, sex, hypertension, diabetes mellitus, smoking status, total cholesterol, systolic blood pressure) was 0.681 (baseline model). Further including the CT-derived AVCS increased this to 0.712 (delta of 0.031 [95%CI: −0.001 to 0.082] vs the baseline model), whereas replacing AVCS (which requires CT imaging) with DASSi (derived from simple TTE) resulted in a C-statistic of 0.714 (delta of 0.033 [95%CI: 0.001-0.083] vs the baseline model).

### Simulated power calculations for DASSi-led clinical trial enrollment

Finally, we sought to evaluate the implications of enriching clinical studies of AS progression based on DASSi vs traditional Doppler parameters. Assuming paired comparisons of changes from baseline, we estimated that enriching for individuals with DASSi ≥0.7 (observed annualized progression rate in AV-V_max_ of 0.37 ± 0.29 m/sec/year) versus all-comers with mild or moderate AS (0.22 ± 0.24 m/sec/year) translated into a 41% reduction in the required sample size to show significant progression from baseline. Similarly, when comparing a DASSi-led strategy versus enrolling individuals with AV-V_max_ ≥ 3 m/sec (annualized AV-V_max_ rate of 0.22 ± 0.26 m/sec/year), we estimated a 45% relative reduction in the required sample size.

## DISCUSSION

We demonstrate that an AI digital biomarker of AS severity (DASSi) deployed on baseline single-view two-dimensional echocardiography enables in-depth phenotyping of aortic valve function, structure and biology with prognostic implications for AS progression and clinical outcomes. In an analysis of a clinical trial of patients with mild or moderate AS undergoing serial multimodality imaging with echocardiography, CT imaging, and [18F]NaF PET-CT, we demonstrate correlations between DASSi and AS severity features that are not routinely derived from echocardiography, such as aortic valve calcium burden and microcalcification activity. More importantly, higher DASSi values at baseline are associated with faster hemodynamic and structural progression in AS severity, independent of traditional risk factors and AVA. Notably, a scalable, AI-augmented echocardiography protocol that incorporates DASSi to standard Doppler estimates demonstrates prognostic performance for future AVR comparable to multimodality assessment by both echocardiography and CT.

Our findings suggest the robustness of DASSi as an objective prognostic tool for the severity and progression of AS in a study with protocolized follow-up with multimodality imaging. The head-to-head comparison against interpretable imaging features spanning hemodynamic, structural and biological aortic valve remodeling offers a degree of explainability that extends beyond current AI methods that rely exclusively on heatmaps over existing images and provide no biological correlations. The work demonstrates that the cardiac and valvular remodeling that track the natural history of AS progression are linked to functional and structural phenotypic information encoded in a PLAX view on routine 2D echocardiography, as estimated using DASSi. In fact, our study shows that AI-augmented interpretation of standard echocardiography could provide insights into advanced imaging features, such as CT-derived AVCS,^16^ the use of which may be limited by accessibility and radiation exposure. Critically, given its simplicity, DASSi can be calculated both retrospectively across existing scans, and prospectively adding no time to the acquisition process.

There are key implications of DASSi to both clinical care and novel therapeutic development in AS. First, our analysis in a prospectively enrolled population with protocolized monitoring strongly supports the utility of DASSi as a scalable diagnostic and prognostic biomarker which may represent an additional read-out in the phenotyping and risk stratification of mild or moderate AS. Second, DASSi offers a promising read-out to guide the enrichment of clinical trials studying the efficacy of medical therapies in AS.^8^ AI-augmented interpretation of standard echocardiographic videos may overcome the operator-dependence of Doppler-derived echocardiographic parameters while offering a scalable approach to phenotype function, structure and provide insights into biology through simple and accessible protocols. As shown in our analysis, we estimated DASSi-led enrollment is associated with a 41-45% lower required sample size to show significant disease progression versus enrollment based on Doppler parameters (AV-V_max_). Thus, the continuous and graded association between DASSi severity and progression, as well as its operator-independence and scalability further ensure favorable statistical properties. Third, given its association with multimodality metrics of AS severity progression and a related signal towards higher risk of AVR, DASSi may offer an intermediate surrogate outcome measure in phase II and early phase III trials. Finally, thanks to its simplicity, it may facilitate broader inclusion of participants across centers that have traditionally lacked access to specialized imaging facilities.

## STUDY LIMITATIONS

Certain limitations merit consideration. First, DASSi phenotyping was not successful performed in 10% of the studies. This was due to the view classification software, which is a part of the fully automated pipeline that assigns labels to the acquired views and may disregard videos for which the automated view classifier has limited confidence that they correspond to a PLAX acquisition.^32^ Second, although the imaging follow-up in the study was protocolized, several patients were eventually referred for AVR. We therefore suspect that the eventual censoring of these patients from the imaging progression analysis may have underestimated the true effect size, given the expected progression in their severity features had they not been referred for AVR. Third, SALTIRE-2 was performed in a single tertiary referral center study, and the composition of the study reflects the local demographics of South-East Scotland where the study was conducted. However, our findings are in line with previous validation studies for DASSi across three international and ethnically diverse cohorts.^14^ Given the mechanistic and multimodality focus of this analysis, the clustering of imaging scans in one study site ensured consistency in the protocols which is a prerequisite for precision in the outcome measures of mechanistic investigations.

## CONCLUSIONS

An AI tool directly applied to 2D-echocardiography defines a personalized profile of AS progression that correlates with several functional, structural, and biological features of AS severity that traditionally require access to multimodality imaging. DASSi-enhanced echocardiography may provide an accessible alternative to multimodality imaging in the phenotyping of non-severe AS in clinical practice and as a scalable and affordable precision biomarker in clinical trials.

## DATA AVAILABILITY

An executable or containerized version of the DASSi algorithm is available and can be made accessible for research use by contacting the corresponding author. The SALTIRE-2 trial data were made available by the study’s principal investigator, Dr. Marc Dweck (University of Edinburgh). No transfer of images or videos occurred across sites.

## ABBREVIATIONS

AS: aortic stenosis
AV-Vmax: peak aortic valve velocity
AVCS: aortic valve calcium score
AVR: aortic valve replacement
CMR: cardiac magnetic resonance imaging
CT: computed tomography
DASSi: Digital Aortic Stenosis Severity Index
PET-CT: positron emission tomography - computed tomography
PLAX: parasternal long axis
RCT: randomized clinical trials
TBR: target-to-background ratio

