## Supplemental Material for "Artificial intelligence-enabled echocardiography as a surrogate for multi-modality aortic stenosis imaging: post-hoc analysis of a clinical trial"

**Online Supplement**

Evangelos K. Oikonomou, MD, DPhil,[a,b] Neil J. Craig, MBChB,[c] Gregory Holste, MSE,[b,d]
Sumukh Vasisht Shankar, MS,[a,b] Audrey White, BSc,[c] Menaka Mahendran, MD,[c]
David E. Newby, MD, PhD,[c] Marc R. Dweck, MD, PhD,[c] Rohan Khera, MD, MS[a,b,e,f*]

[a] Section of Cardiovascular Medicine, Dept of Internal Medicine, Yale School of Medicine, New Haven, CT, USA
[b] Cardiovascular Data Science (CarDS) Lab, Yale School of Medicine, New Haven, CT, USA
[c] Centre for Cardiovascular Science, University of Edinburgh, Edinburgh, United Kingdom
[d] Department of Electrical and Computer Engineering, University of Texas in Austin, Austin, TX, USA
[e] Section of Biomedical Informatics and Data Science, Yale School of Medicine, New Haven, CT, USA
[f] Section of Health Informatics, Department of Biostatistics, Yale School of Public Health, New Haven, CT, USA

**Table of contents:**
**Supplemental Table 1** – page 2
**Supplemental Figure 1** – page 3

***Address for correspondence:**
Rohan Khera, MD, MS
195 Church St, 6th Floor, New Haven, CT 06510
203-764-5885;

**Supplemental Table 1 | Association between DASSi and cross-sectional aortic valve calcification and calcification activity.**

| Modality | Dependent variable | Variable | Coefficient | 95% CI Lower | 95% CI Upper | *p* value |
|---|---|---|---|---|---|---|
| TTE | **AVA (cm$^2$)** | DASSi* | -0.07 | -0.12 | -0.02 | 0.003 |
| | **AV-$V_{max}$ (m/sec)** | DASSi* | 0.21 | 0.12 | 0.30 | <0.001 |
| | **AV mean gradient (mmHg)** | DASSi* | 3.68 | 2.18 | 5.19 | <0.001 |
| CT | **AVCS** | AVA (cm$^2$) | -730.28 | -1358.85 | -101.70 | 0.023 |
| | | DASSi* | 283.70 | 100.80 | 466.60 | 0.002 |
| PET-CT | **[18F]NaF $TBR_{max}$** | AVA (cm$^2$) | -0.38 | -0.84 | 0.07 | 0.10 |
| | | DASSi* | 0.17 | 0.04 | 0.31 | 0.011 |
| | **[18F]NaF $TBR_{mean}$** | AVA (cm$^2$) | -0.20 | -0.38 | -0.03 | 0.023 |
| | | DASSi* | 0.08 | 0.03 | 0.13 | 0.004 |

* DASSi coefficients refer to one standard deviation increments. The estimates are derived at the baseline visit from a generalized linear model with baseline DASSi, age, sex, hypertension, diabetes, smoking status, total cholesterol, systolic blood pressure as independent variables. For CT and PET-CT analysis, AVA is included as an additional independent predictor. *AV: aortic valve; AV-$V_{max}$: peak aortic valve velocity; AVA: aortic valve area; AVCS: aortic valve calcium score; CI: confidence interval; CT: computed tomography; DASSi: digital aortic stenosis severity index; NaF: sodium fluoride; PET: positron emission tomography; TBR: target-to-background ratio; TTE: transthoracic echocardiography.*

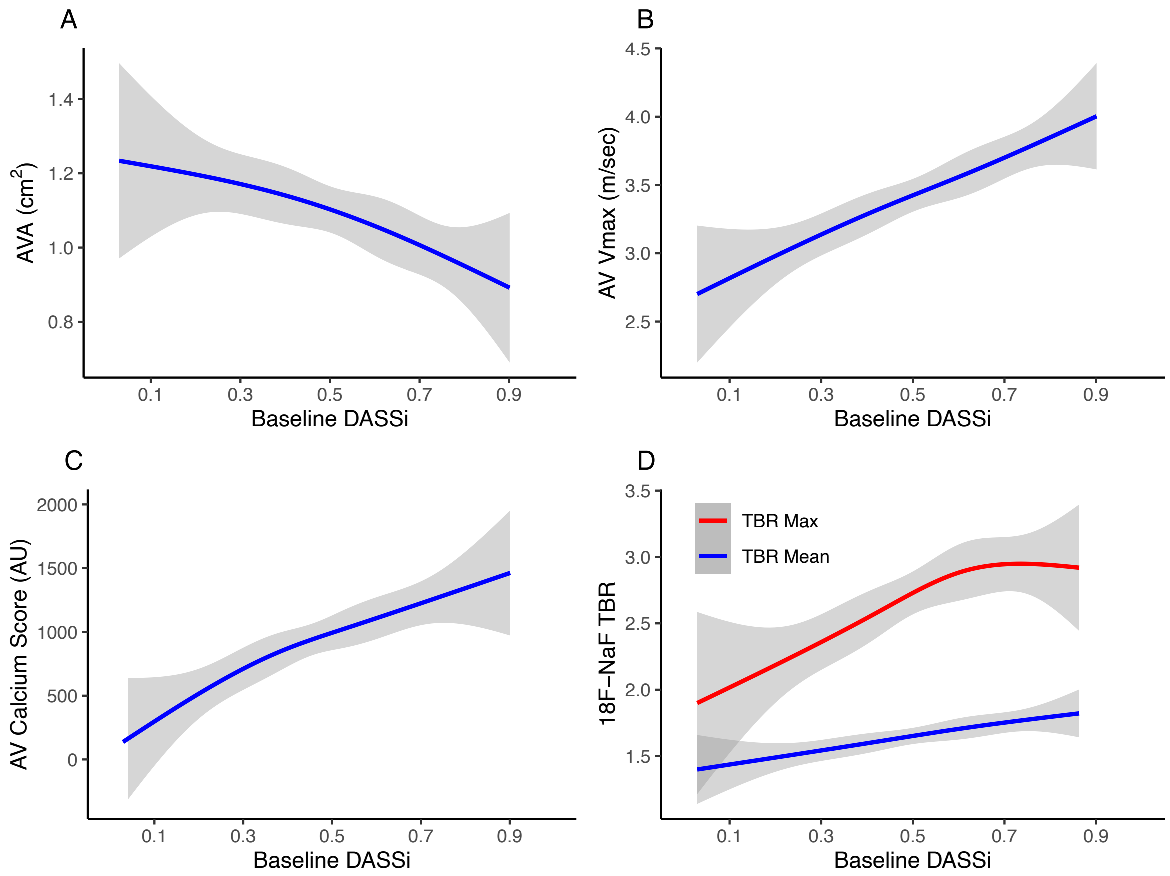


**Supplemental Figure 1 | Cross-sectional correlation of DASSi with multimodality AS phenotyping.** Adjusted spline curves (with 3 knots) illustrating the cross-sectional association between DASSi on the x axis, and (**A**) AVA, (**B**) AV-$V_{max}$, (**C**) CT-derived aortic valve calcium score (AVCS); (**D**) [18F]NaF PET-CT-derived $TBR_{mean}$ and $TBR_{max}$ at the level of the aortic valve. *AS: aortic stenosis; AV: aortic valve; AV-$V_{max}$: peak aortic valve velocity; AVA: aortic valve area; AVCS: aortic valve calcium score (AVCS); AU: Agatston Units; CT: computed tomography; DASSi: digital aortic stenosis severity index; NaF: sodium fluoride; PET: positron emission tomography; TBR: target-to-background ratio.*
